# Determinants of in-hospital mortality within 48 hours of admission to the Emergency and Urgent Care Department at University Teaching Hospital, Lusaka, Zambia: a retrospective cross-sectional study

**DOI:** 10.64898/2026.05.07.26352696

**Authors:** Joop Tambo

## Abstract

**Background:** The emergency department (ED) serves as a critical entry point into hospital care and a sentinel indicator of health system performance. In-hospital mortality within 48 hours of ED admission represents acute care failures that are often preventable yet remain poorly characterized in sub-Saharan African (SSA) settings. This study aimed to identify the demographic, clinical, and hospital-related determinants of in-hospital mortality within 48 hours of admission to the Emergency and Urgent Care Department at the University Teaching Hospital (UTH), Lusaka, Zambia.

**Methods:** A retrospective cross-sectional study was conducted using 385 patient records from UTH’s Emergency and Urgent Care Department for the year 2021. Data were extracted from the District Health Information System 2 (DHIS2) using simple random sampling. Descriptive statistics, univariate, and multivariable logistic regression analyses were performed using STATA 16.1 MP. Variables with p<0.20 in univariate analysis were retained for adjusted modelling. Multicollinearity was assessed via variance inflation factors (VIF <5). Model fit was evaluated using the Hosmer-Lemeshow goodness-of-fit test and receiver operating characteristic (ROC) analysis.

**Results:** Of 385 patients, 175 (45.5%) died within 48 hours of admission. Patients who died were older (median age 45 vs. 37.5 years, p<0.001). In multivariable analysis, three variables were independently associated with 48-hour mortality: pulse rate (aOR = 0.98, 95% CI: 0.95–1.00, p = 0.036), Glasgow Coma Scale (GCS) score (aOR = 0.75, 95% CI: 0.63–0.90, p = 0.002), and out-of-hours admission between 00:00–07:59 (aOR = 11.44, 95% CI: 1.19–109.96, p = 0.035). Age was a significant predictor in univariate analysis but not in the adjusted model, indicating confounding. The model demonstrated good discriminatory ability (AUC = 0.81).

**Conclusions:** Reduced pulse rate, lower GCS score at admission, and out-of-hours presentation are independent determinants of 48-hour in-hospital mortality at UTH. These findings underscore the need for enhanced vital sign monitoring protocols, targeted staffing during overnight hours, and improved risk stratification tools in resource-constrained emergency care settings. The wide confidence interval for the time-of-admission finding warrants cautious interpretation and validation in future prospective studies.

## Introduction

The emergency department (ED) functions as the critical interface between communities and hospital-based care, providing urgent attention to acutely ill and injured patients. Beyond its clinical role, ED performance serves as a sentinel indicator of broader health system capacity and quality [1]. Globally, in-hospital mortality within 48 hours of ED admission is recognized as an acute care quality measure that captures immediate failures in stabilization, triage, and patient management—failures that are disproportionately common in low- and middle-income countries (LMICs) [2,3].

The World Health Organization estimates that effective emergency care systems could prevent up to 50% of deaths in LMICs, where mortality rates remain significantly higher than in high-income settings [4]. In sub-Saharan Africa (SSA), early in-hospital mortality is compounded by structural barriers including inadequate staffing, diagnostic resource limitations, delayed referrals, and a dual burden of communicable and non-communicable disease [5,6]. Studies from the region consistently identify vital sign derangements, level of consciousness, and out-of-hours admission as key contributors to early mortality, yet most evidence is drawn from East African contexts, with limited data from Zambia specifically [7,8].

Zambia’s National Health Strategic Plan (NHSP) 2022–2026 sets an explicit target of reducing 48-hour in-hospital mortality to 3 per 1,000 admissions [9]. The University Teaching Hospital (UTH) in Lusaka, the nation’s sole quaternary referral centre and largest tertiary care facility, bears a disproportionate burden of acute and emergency admissions nationally. Despite its centrality to emergency care delivery, the specific determinants of early mortality at UTH remain poorly characterized. Existing UTH data are limited to disease-specific cohorts—such as sepsis [10] and COVID-19 pneumonia [11]—and do not provide a comprehensive picture of 48-hour mortality determinants across the full range of ED presentations.

This study addresses that gap by investigating the demographic, clinical, and hospital-level determinants of in-hospital mortality within 48 hours of admission to UTH’s Emergency and Urgent Care Department. By identifying modifiable predictors across medical and surgical admissions during 2021—a period characterized by the COVID-19 pandemic—this research aims to inform targeted interventions, risk stratification protocols, and staffing policies to reduce preventable early deaths in this resource-limited setting.

## Methods

### Study design and setting

A retrospective cross-sectional study was conducted at the Emergency and Urgent Care Department (EUCD) of University Teaching Hospital–Adult (UTH-A), Lusaka, Zambia. UTH is the largest tertiary and quaternary referral facility in Zambia, serving a catchment population from across the country. The EUCD operates 24 hours a day and manages both medical and surgical emergency admissions. The study period covered all admissions from 1 January to 31 December 2021, during which Zambia experienced the third and fourth waves of the COVID-19 pandemic.

### Study population and sampling

The sampling frame comprised all patients admitted to the EUCD during the study period, as captured in the District Health Information System 2 (DHIS2). The minimum required sample size of 385 was calculated using the formula n = Z^2^p(1−p)/e^2^, assuming a conservative prevalence estimate of 50% (p = 0.50), a 95% confidence level (Z = 1.96), and a margin of error of 5% (e = 0.05). Simple random sampling was applied to select patient records from the DHIS2 database, ensuring representativeness.

### Inclusion and exclusion criteria

Patients were eligible if they were admitted to the EUCD between January and December 2021, and if their outcome (death within 48 hours or survival beyond 48 hours) was documented. Records were excluded if more than 50% of key variables—including vital signs or outcome data—were missing.

### Variables

The primary outcome was in-hospital mortality within 48 hours of EUCD admission, defined as death occurring from the time of ED registration until 48 hours thereafter, regardless of the underlying cause, as documented in the medical record.

Explanatory variables were categorised as follows: (i) demographic factors—age (years) and sex (male/female); (ii) clinical factors—reason for admission (respiratory vs. non-respiratory), coexisting conditions (classified per ICD-11), oxygen saturation (SpO_2_, %), systolic blood pressure (mmHg), diastolic blood pressure (mmHg), pulse rate (beats per minute), body temperature (°C), and level of consciousness assessed using the Glasgow Coma Scale (GCS, scored 3–15); and (iii) hospital factors—day of admission (weekday vs. weekend), time of admission (00:00–07:59 vs. 08:00–23:59), type of admission unit (medical vs. surgical), and level of referring facility (tiered 1–4, health centre, self-referral/private).

### Data management and statistical analysis

Data were extracted from patient medical records and DHIS2, validated against original files, and coded using dummy variables as appropriate. Missing data for continuous variables were handled via listwise deletion in multivariable models; sensitivity analyses using mean imputation confirmed the robustness of findings.

Continuous variables were assessed for normality using the Shapiro–Wilk test and reported as mean (±SD) or median [interquartile range (IQR)] accordingly. Categorical variables were summarized as frequencies and proportions. Bivariate comparisons between decedents and survivors used the Mann–Whitney U test for non-normally distributed continuous variables and the chi-squared test (or Fisher’s exact test where expected cell counts were <5) for categorical variables.

Multivariable logistic regression was used to model the binary outcome. An investigator-led backward stepwise approach was employed, retaining variables with p<0.20 in univariate analysis. Multicollinearity was assessed using variance inflation factors (VIF), with a threshold of VIF <5. Model performance was evaluated using the Hosmer–Lemeshow goodness-of-fit test and the area under the receiver operating characteristic (ROC) curve (AUC). Results are reported as crude odds ratios (cOR) and adjusted odds ratios (aOR) with 95% confidence intervals (CIs). Statistical significance was set at p<0.05. All analyses were performed using STATA 16.1 MP (StataCorp, College Station, TX, USA).

### Ethics

Ethics approval was granted by the University of Zambia Biomedical Research Ethics Committee (UNZA-BREC REF 3471-2022) and by the National Health Research Authority (NHRA). Further institutional approval was obtained from the UTH Senior Medical Superintendent. Patient confidentiality was maintained throughout; all data were anonymised prior to analysis.

## Results

### Patient characteristics

A total of 385 patient records were included in the analysis, of whom 175 (45.5%) died within 48 hours of admission and 210 (54.5%) survived beyond 48 hours. The median age of all patients was 40 years (IQR: 27–61). Patients who died within 48 hours were significantly older (median 45 years, IQR: 29–67) compared with survivors (median 37.5 years, IQR: 22.5–56.5; p<0.001). The majority of patients were male (62.3%; n = 240). Sex was not significantly associated with 48-hour mortality (p = 0.409). Sociodemographic characteristics are presented in Table 1.

**Table 1.** Sociodemographic and clinical characteristics of patients admitted to the Emergency and Urgent Care Department, UTH, 2021.

| Characteristic | Overall $n=385$<br>$n$ (%) | Deaths $\leq 48$hrs<br>$n=175$ $n$ (%) | Survived $>48$hrs<br>$n=210$ $n$ (%) | p-value |
| --- | --- | --- | --- | --- |
| Age, years; median (IQR) | 40 (27–61) | 45 (29–67) | 37.5 (22.5–56.5) | $<0.001$ U |
| Sex: Male | 240 (62.3) | 113 (64.6) | 127 (60.5) | 0.409 C |
| Sex: Female | 145 (37.7) | 62 (35.4) | 83 (39.5) |  |
| Reason for admission<br>($n=378$) | | | | |
| Respiratory | 77 (20.4) | 45 (25.9) | 32 (15.7) | 0.014 C |
| Non-respiratory | 301 (79.6) | 129 (74.1) | 172 (84.3) |  |
| Oxygen saturation<br>($n=292$); median (IQR) | 95 (90–97) | 93 (83–97) | 96 (94–98) | $<0.0001$ U |
| Pulse rate ($n=344$);<br>median (IQR) | 102 (87–119) | 104 (89–122) | 100 (86–116) | 0.234 U |
| GCS score ($n=147$);<br>median (IQR) | 11 (8–15) | 10.5 (7–15) | 15 (10–15) | 0.003 U |
| Systolic BP ($n=343$);<br>median (IQR) | 127 (110–147) | 130.5 (111.5–<br>148.5) | 124 (107–143) | 0.132 U |
| Diastolic BP ($n=343$);<br>median (IQR) | 77 (61–88) | 78 (60–90) | 75 (63–87) | 0.762 U |
| Body temperature<br>($n=217$); median (IQR) | 36.2 (35.9–<br>36.6) | 36.4 (35.8–36.7) | 36.2 (35.9–36.5) | 0.212 U |

U = Mann-Whitney U test; C = Chi-squared test; BP = blood pressure; GCS = Glasgow Coma Scale; IQR = interquartile range. Bold p-values denote statistical significance ( $p < 0.05$ ).

### Hospital characteristics

The majority of admissions occurred on weekdays (73.0%; n = 281). Notably, most patients (86.5%; n = 326) were admitted between 00:00 and 07:59 hours. Medical unit admissions constituted 62.9% (n = 242) of the total. Mortality within 48 hours was significantly higher among medical unit admissions compared with surgical admissions (75.4% vs. 24.6%, p<0.0001 by chi-squared). The largest proportion of patients were referred from first-level hospitals (44.7%). Hospital characteristics are presented in Table 2.

**Table 2.** Hospital characteristics of patients admitted to the Emergency and Urgent Care Department, UTH, 2021.

| Characteristic | Overall $n=385$<br>$n$ (%) | Deaths $\leq 48$hrs<br>$n=175$ $n$ (%) | Survived $>48$hrs<br>$n=210$ $n$ (%) | p-value |
| --- | --- | --- | --- | --- |
| Day of admission |  |  |  |  |
| Weekday | 281 (73.0) | 121 (69.1) | 160 (76.2) | 0.121 C |
| Weekend | 104 (27.0) | 54 (30.9) | 50 (23.8) |  |
| Time of admission |  |  |  |  |
| 00:00–07:59 | 326 (86.5) | 146 (83.9) | 180 (88.7) | 0.178 C |
| 08:00–23:59 | 51 (13.5) | 28 (16.1) | 23 (11.3) |  |
| Admission unit |  |  |  |  |
| Medical | 242 (62.9) | 132 (75.4) | 110 (52.4) | $<0.0001$ C |
| Surgical | 143 (37.1) | 43 (24.6) | 100 (47.6) |  |
| Referring facility level |  |  |  |  |
| Not recorded | 85 (22.1) | 50 (28.6) | 35 (16.7) | 0.058 C |
| Level 1 hospital | 172 (44.7) | 68 (38.9) | 104 (49.5) |  |
| Level 2 hospital | 34 (8.8) | 13 (7.4) | 21 (10.0) |  |
| Level 3 hospital | 3 (0.8) | 1 (0.6) | 2 (1.0) |  |
| Health centre | 74 (19.2) | 33 (18.9) | 41 (19.5) |  |
| Self-referral/private | 17 (4.4) | 10 (5.7) | 7 (3.3) |  |

C = Chi-squared test.

### Multivariable analysis

Results of the crude and adjusted logistic regression analyses are presented in Table 3. In the adjusted model, three variables were independently associated with 48-hour in-hospital mortality. Pulse rate was inversely associated with mortality (aOR = 0.98, 95% CI: 0.95–1.00, p = 0.036), indicating that each one-unit increase in pulse rate was associated with a 2% reduction in the odds of death. This pattern may reflect the higher pulse rates observed among survivors as a physiological compensatory response.

**Table 3.** Crude and adjusted odds of 48-hour in-hospital mortality among patients admitted to the Emergency and Urgent Care Department, UTH, 2021.

| Variable | cOR | 95% CI | p-value | aOR | 95% CI | p-value |
| --- | --- | --- | --- | --- | --- | --- |
| Age (per year) | 1.02 | 1.01–1.03 | 0.001* | 1.01 | 0.98–1.04 | 0.655 |
| Sex: Female (ref: Male) | 0.84 | 0.55–1.27 | 0.409 | 1.98 | 0.66–5.94 | 0.225 |
| Reason: Non-respiratory (ref: Respiratory) | 0.53 | 0.32–0.89 | 0.015* | 0.64 | 0.16–2.60 | 0.533 |
| Oxygen saturation | 0.94 | 0.91–0.96 | <0.0001* | 0.97 | 0.93–1.01 | 0.124 |
| Systolic BP | 1.00 | 1.00–1.01 | 0.211 | 0.98 | 0.96–1.01 | 0.168 |
| Diastolic BP | 1.00 | 0.99–1.01 | 0.897 | 0.98 | 0.94–1.01 | 0.262 |
| Pulse rate | 1.00 | 0.99–1.01 | 0.471 | 0.98 | 0.95–1.00 | 0.036* |
| GCS score | 0.87 | 0.79–0.96 | 0.004* | 0.75 | 0.63–0.90 | 0.002* |
| Weekend (ref: Weekday) | 1.43 | 0.91–2.24 | 0.122 | 2.19 | 0.58–8.19 | 0.246 |
| 00:00–07:59 (ref: 08:00–23:59) | 1.50 | 0.83–2.72 | 0.180 | 11.44 | 1.19–109.96 | 0.035* |
| Surgical unit (ref: Medical) | 0.36 | 0.23–0.56 | <0.0001* | 0.87 | 0.20–3.77 | 0.847 |

aOR = adjusted odds ratio; cOR = crude odds ratio; CI = confidence interval; BP = blood pressure; GCS = Glasgow Coma Scale. \* $p < 0.05$ .

Higher GCS score at admission was independently protective against mortality (aOR = 0.75, 95% CI: 0.63–0.90, p = 0.002), such that each one-point increase in GCS was associated with a 25% reduction in the adjusted odds of death. This association was also significant in the unadjusted model (cOR = 0.87, 95% CI: 0.79–0.96, p = 0.004).

Admission between 00:00 and 07:59 was associated with substantially higher odds of mortality compared to admission between 08:00 and 23:59 (aOR = 11.44, 95% CI: 1.19–109.96, p = 0.035). This finding was not statistically significant in the unadjusted model (cOR = 1.50, p = 0.180), suggesting that the effect of out-of-hours admission emerges after adjustment for case mix. The wide confidence interval reflects sparse data in the reference category and should be interpreted with appropriate caution.

Age was significantly associated with mortality in the unadjusted analysis (cOR = 1.02, 95% CI: 1.01–1.03, p = 0.001) but was not retained as a significant predictor in the adjusted model (aOR = 1.01, 95% CI: 0.98–1.04, p = 0.655), indicating confounding by clinical variables. Sex, reason for admission, oxygen saturation, blood pressure, body temperature, admission unit, weekend admission, and level of referring facility were not independently associated with 48-hour mortality in the adjusted model.

#### Model diagnostics

The Hosmer–Lemeshow goodness-of-fit test indicated adequate model fit (p = 0.45). The model demonstrated good discriminatory performance, with an AUC of 0.806 (95% CI not reported; sensitivity 72%, specificity 78%), indicating that the model correctly classifies approximately 80.6% of cases. This is substantially above chance and supports the clinical utility of the identified predictors for risk stratification.

## Discussion

This study is among the first to systematically characterise the determinants of 48-hour in-hospital mortality across the full spectrum of emergency admissions at a quaternary referral centre in Zambia. Using data from 385 patients admitted during the COVID-19-affected year of 2021, we identified three independent predictors of early mortality: pulse rate, GCS score at admission, and out-of-hours admission time. These findings carry direct implications for emergency care quality improvement in resource-constrained settings.

The significant association between lower GCS scores and increased mortality is consistent with extensive literature from both high-income and SSA settings. Low GCS has been identified as a predictor of paediatric and adult mortality in emergency settings across the region [12,13], reflecting the downstream consequences of delayed presentation, undertreated sepsis, or traumatic brain injury. In our cohort, the median GCS among decedents was 10.5 compared with 15 among survivors, a clinically important differential. The GCS is a low-cost, universally applicable assessment tool; our findings reinforce calls for its systematic use as a triage criterion at first contact in African emergency departments [14].

The inverse association between pulse rate and mortality in the adjusted model (aOR = 0.98, p = 0.036) is clinically interpretable as a reflection of compensatory physiology: patients who survived were more likely to be maintaining adequate cardiovascular response, whereas those who died may have progressed to cardiovascular decompensation. This pattern is broadly consistent with existing evidence linking abnormal vital sign composites with deterioration [15], though the marginal magnitude of the per-unit odds ratio underscores the importance of considering vital signs as dynamic composite patterns rather than isolated values.

The strong association between out-of-hours admission (00:00–07:59) and increased mortality (aOR = 11.44, p = 0.035) is a critical finding. This period is typically characterized by reduced staffing levels, limited diagnostic capacity, and delayed specialist review—factors consistently linked to worse outcomes in emergency departments globally [16]. In SSA specifically, out-of-hours effects are amplified by structural vulnerabilities, including absent covering specialists and inadequate supervision of junior staff [7]. Notably, this association only emerged in the adjusted model, suggesting that case-mix confounding (e.g., sicker patients presenting overnight) may mask the staffing effect when unadjusted. While the wide confidence interval (1.19–109.96) cautions against over-interpretation—likely reflecting the small number of events in the reference category—the direction and significance of the effect are consistent with prior evidence and warrant urgent policy attention [16,17].

Age was a significant predictor in univariate analysis but attenuated to non-significance in the adjusted model, suggesting that its effect is mediated or confounded by clinical parameters such as GCS and oxygen saturation. This finding is consistent with Porcel-Gálvez et al. (2020) [18], who similarly reported age-associated mortality in univariate but not multivariable analyses in an acute care hospital study. In SSA contexts, the age-mortality relationship may be particularly susceptible to confounding by late presentation and comorbidity burden. The high prevalence of retroviral disease (39.2%) and hypertension (37.1%) among patients with documented comorbidities in our sample reflects the dual epidemic confronting Zambia’s healthcare system, though neither reached statistical significance as an independent predictor, possibly due to the relatively small proportion with complete comorbidity records.

Mortality was significantly higher among medical unit admissions in univariate analysis (75.4% of deaths vs. 24.6% from surgical), consistent with evidence from Spain [18] and other SSA settings [19]. Medical admissions tend to include patients with sepsis, respiratory failure, and advanced organ dysfunction—conditions associated with high short-term fatality in resource-limited EDs. The attenuation of this effect in the adjusted model suggests that clinical parameters, particularly GCS and pulse, explain much of the medical–surgical mortality differential.

### Strengths and limitations

This study has several notable strengths. It is, to our knowledge, the first comprehensive investigation of 48-hour mortality determinants across all emergency admissions at UTH, drawing on a robust sample collected during the COVID-19 pandemic period, which enhances generalizability to similar LMIC emergency settings operating under resource stress. The use of a validated logistic regression framework, STROBE-compliant reporting, and model diagnostic testing add to the rigor of the analysis.

Several limitations must be acknowledged. The retrospective design relies on the completeness of routine medical records; missing data for key variables—particularly GCS (n = 147/385; 38.2%) and body temperature (n = 217/385; 56.4%)—may have reduced statistical power and introduced information bias, though sensitivity analyses using mean imputation produced broadly similar results. The study is restricted to a single tertiary hospital, limiting direct generalizability to lower-level facilities where emergency care capacity differs substantially. Selection bias due to referral patterns cannot be excluded. Additionally, the wide confidence interval for the out-of-hours admission odds ratio reflects sparse data in the reference category and should be interpreted cautiously; prospective validation with larger sample sizes is required. Potentially important predictors such as nurse-to-patient ratios and time from triage to intervention were not available in the routine data and could not be assessed.

## Conclusions

This study identifies pulse rate, GCS score at admission, and out-of-hours presentation as independent determinants of in-hospital mortality within 48 hours at Zambia’s University Teaching Hospital. These findings have immediate clinical and policy relevance. First, systematic GCS assessment at triage should be standardized as a core component of emergency triage protocols. Second, the mortality risk associated with overnight admissions demands specific, policy-driven staffing improvements during these hours—at a minimum, achieving recommended nurse-to-patient ratios and ensuring the availability of supervising clinicians. Third, composite vital sign monitoring tools such as the National Early Warning Score (NEWS) should be evaluated and validated for the UTH context to improve real-time risk stratification.

The attenuated role of age and unit type in adjusted models reinforces the primacy of modifiable clinical and operational factors over non-modifiable demographic characteristics in explaining early mortality. Future prospective studies with complete vital sign data, larger samples, and inclusion of process variables (e.g., time to first intervention, staffing ratios) are needed to deepen understanding of the pathways linking structural and clinical determinants of emergency mortality in Zambia and the broader SSA region.

## Supporting Information

S1 Checklist. STROBE checklist for cross-sectional studies.

S1 Data. An anonymized dataset underlying this study. Available upon reasonable requests from the corresponding author, subject to UTH institutional data governance requirements.

## Acknowledgements

The authors thank the University Teaching Hospital-Adult for providing access to secondary data, and Dr. Soka Nyirenda (UTH-Adult) for valuable discussions. The supervisory contributions of Dr. Jerry Banda and Mrs. Mwiche Musukuma Thole during the primary study are gratefully acknowledged.

## Data Availability

The anonymized dataset underlying this study is available upon reasonable request to the corresponding author, subject to institutional data governance requirements at UTH. The data were sourced from hospital medical records and the DHIS2 system and are not publicly deposited due to patient confidentiality obligations.

---

**Figure 1.**
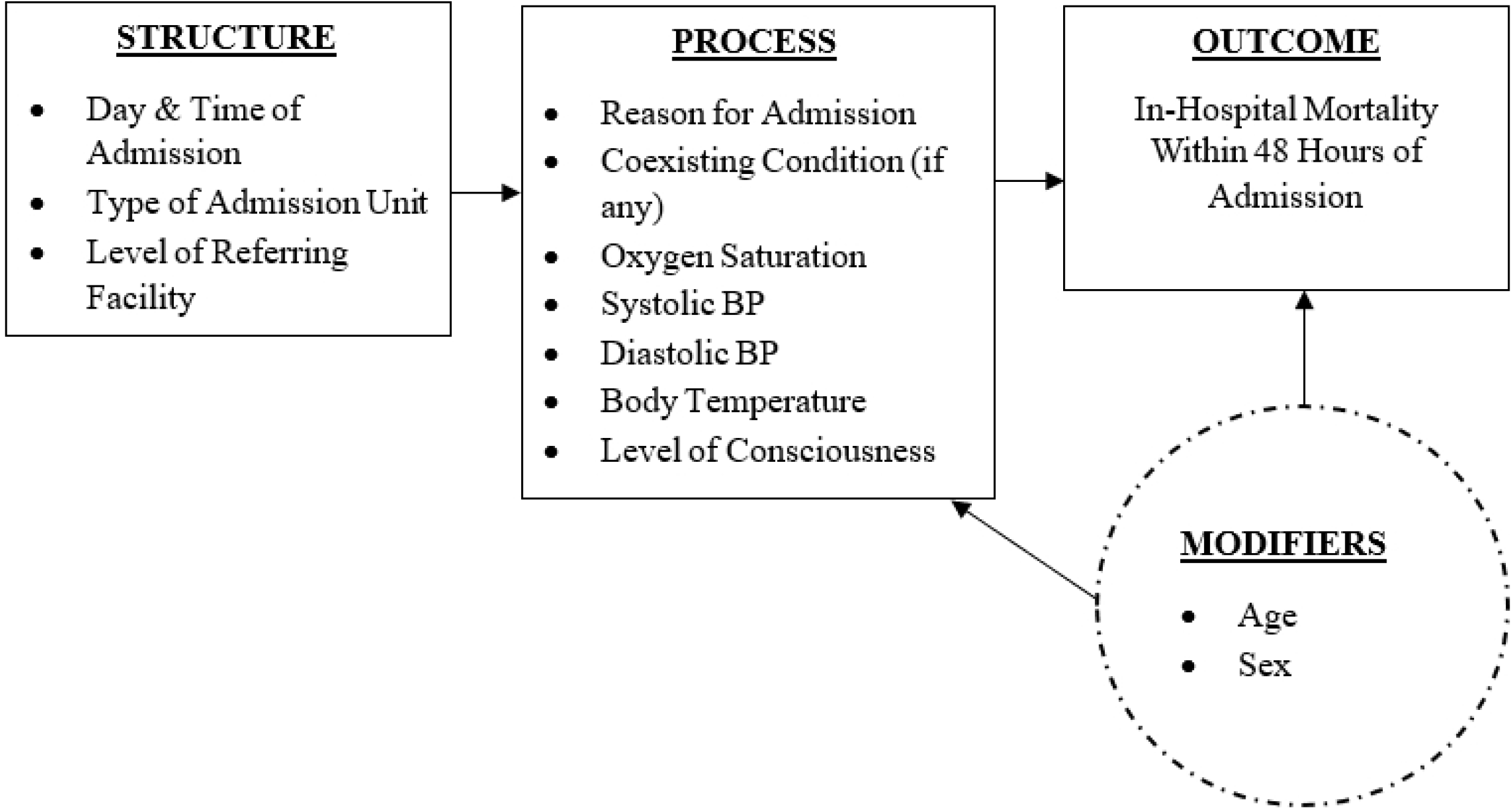
Conceptual Framework Based on the Donabedian Model. Conceptual Framework

**Figure 2:**
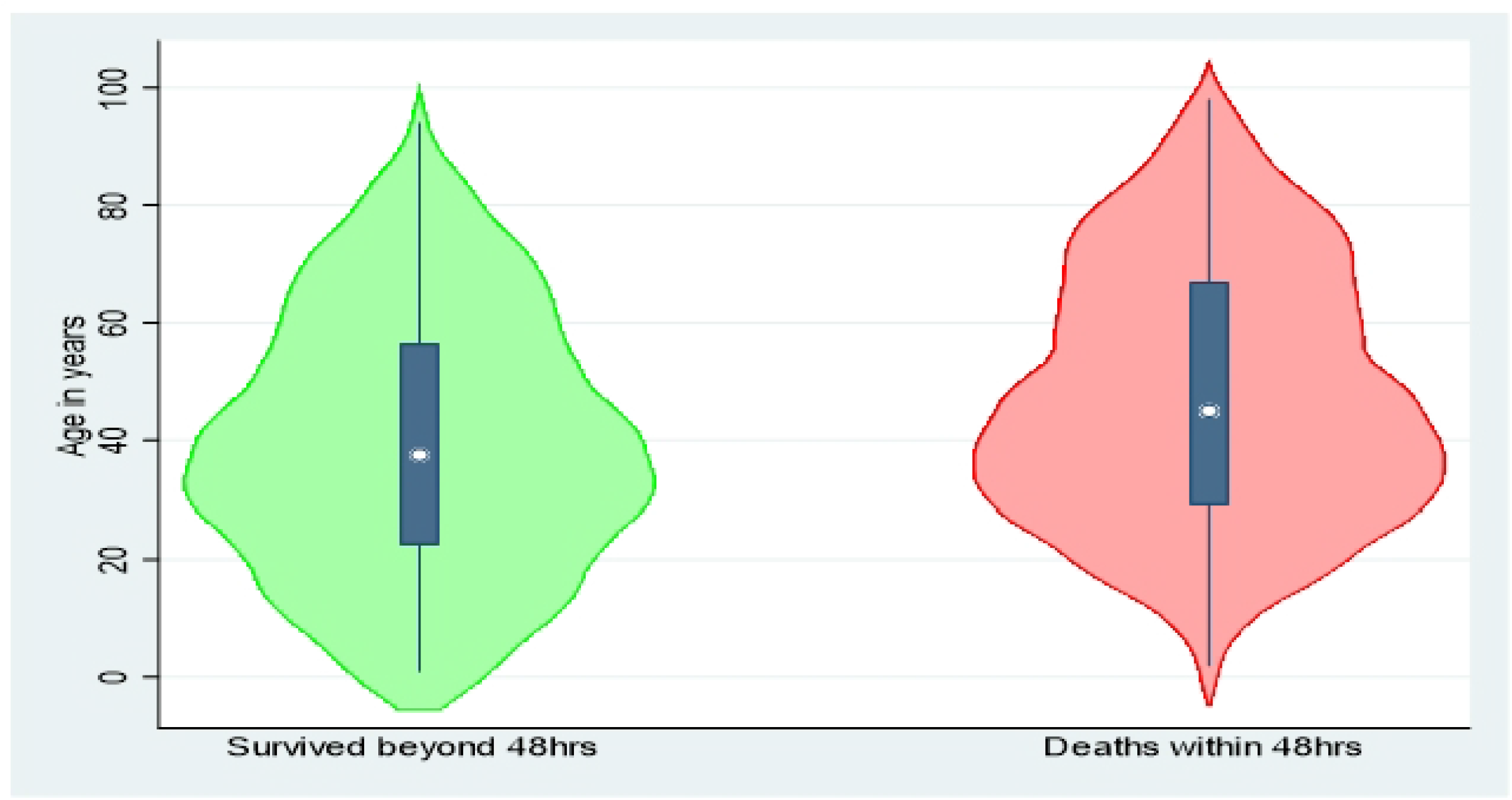
Age vs Outcome of in-hospital mortalities within 48hours of admission. Age vs Outcome

**Figure 3.**
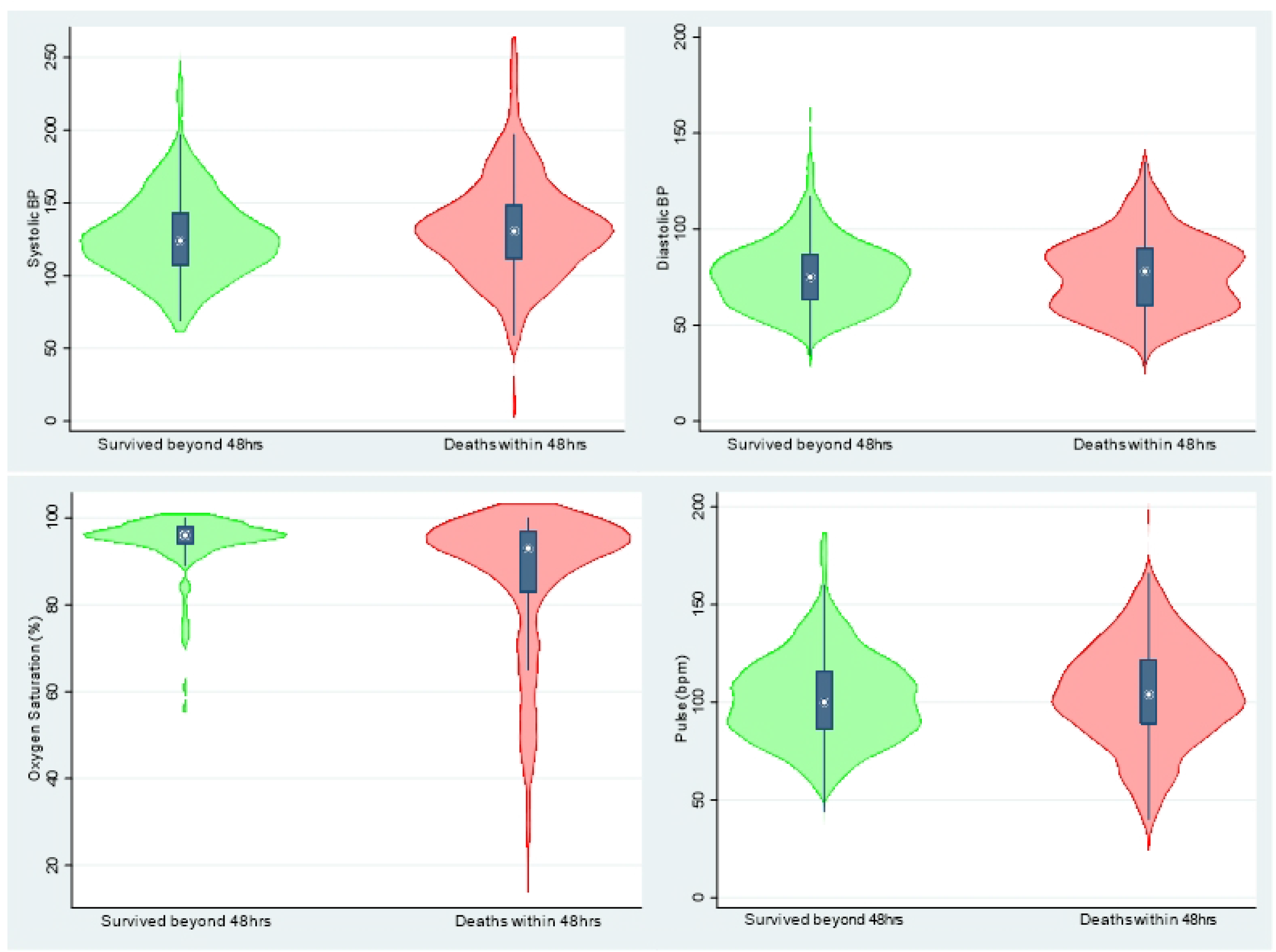
Vital statistics vs. outcome of in-hospital mortalities within 48hours of admission. Vital Stats vs Outcome

**Figure 4.**
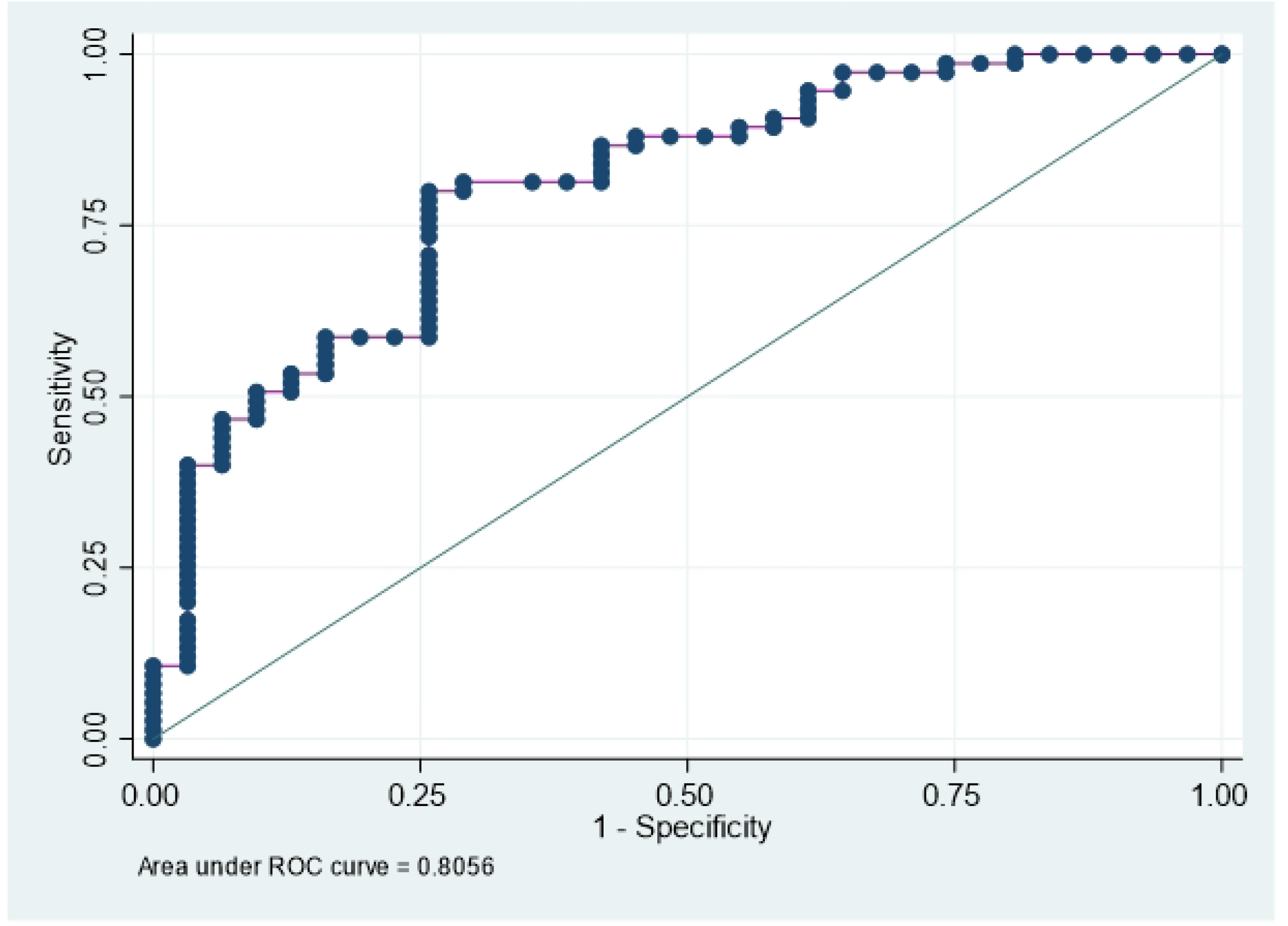
ROC curve for in-hospital mortalities within 48hrs for the identified determinants. ROC Curve

